# Extracellular Microenvironment in Patient-derived Hydrogel Organoids of Prostate Cancer Regulates Therapeutic Response

**DOI:** 10.1101/2020.05.17.20104349

**Authors:** Matthew J Mosquera, Rohan Bareja, Jacob Bernheim, Muhammad Asad, Cynthia Cheung, Michael Sigouros, Varun Prabhu, Joshua E Allen, M. Laura Martin, Loredana Puca, Mark Rubin, Himisha Beltran, Juan Miguel Mosquera, Olivier Elemento, Ankur Singh

## Abstract

Following treatment with androgen receptor (AR) pathway inhibitors, ~20% of prostate cancer patients progress by shedding their dependence on AR. These tumors undergo epigenetic reprogramming turning castration-resistant prostate cancer adenocarcinoma (CRPC-Adeno) into neuroendocrine prostate cancer (CRPC-NEPC). Currently, no targeted therapies are available for CRPC-NEPCs. A major hurdle in the development of new therapies and treatment of CRPC-NEPC is the lack of accurate models to test candidate treatments. Such models would ideally capture components of the tumor microenvironment (TME) factors, which likely regulate the phenotypic, genetic, and epigenetic underpinnings of this aggressive subset. The TME is a complex system comprised not only of malignant prostate cells but also stromal and inflammatory cells and a scaffold of extracellular matrix (ECM). ECM proteins are implicated in the survival and progression of cancer and development of chemoresistance, while are equally integral to the development of prostate cancer organoids. Here, using a combination of patient tumor proteomics and RNA sequencing, we define putative ECM cues that may guide the growth of prostate tumors in patients. Using this molecular information, we developed synthetic hydrogels that recapitulate the tumor ECM. Organoids cultured in the synthetic hydrogel niches demonstrate that ECM subtypes regulate the morphology, transcriptome, and epigenetics hallmarks of CRPC-Adeno and CRPC-NEPC. CRPC-NEPC organoid showed a differential response to small molecule inhibitors of epigenetic repressor EZH2 and Dopamine Receptor D2 (DRD2), the latter being a novel target in CRPC-NEPC when grown in tumor-specific ECM. Finally, in those synthetic ECM niches where drug resistance was observed in CRPC-NEPCs, cellular reprogramming by a synergistic combination of EZH2 inhibitors with DRD2 antagonists inhibited tumor growth. The synthetic platform can provide a more realistic prostate-specific microenvironment and subsequently enable the development of effective targeted therapeutics for prostate cancers.

## Main

The acquired drug resistance in advanced prostate cancer to current therapies, including next-generation AR pathway inhibitors, such as abiraterone acetate and enzalutamide, is driven, in part, by the ability of cancer cells to adopt AR-independent pathways for growth and survival^1-3^. Emerging evidence suggests that prolonged AR pathway inhibition can alter the archetypical course of the disease, leading to treatment-induced lineage transition where cellular dedifferentiation and alterations in the lineage of prostate cancer cells in the form of epithelial-mesenchymal transition (EMT) and/or neuroendocrine differentiation^1, 4–6^. A highly aggressive and lethal form of this lineage plasticity is when castration-resistant adenocarcinoma (CRPC-Adeno) tumors develop clinical transformation to a small cell neuroendocrine carcinoma-like presentation, termed CRPC-NEPC^7^, which manifests heterogenous histological features^8^.

A prevailing hypothesis suggests that CRPC-NEPCs evolves from CRPC-Adeno through lineage conversion to the neuroendocrine lineage via genetic and/or dysregulation of at least one epigenetic modifier, histone methyltransferase enhancer of zeste 2 (EZH2)^7, 9–12^. Patients with CRPC-NEPC do not tend to respond well to standard drugs used to treat CRPC and are often treated with platinum-based chemotherapy. However, these tumors typically progress and there are no standard next line options. Currently, no targeted therapies are available for CRPC-NEPC. The limited therapeutic options underscore the need for more sophisticated modeling of tumors to predict response to standard and new therapies while considering patient-specific microenvironment heterogeneity in which these tumors evolve, progress, undergo transformations, change metastatic behavior, and regulate response to therapeutics.

The lack of *ex vivo* prostate cancer models that recapitulate patient- or disease-specific features of human CRPC-Adeno and CRPC-NEPC has significantly hampered progress in understanding disease pathogenesis and therapy response. To partly overcome this major roadblock, we have recently established patient-derived Matrigel-based organoids of prostate cancer^13^. These organoids retain the genomic and transcriptome features of the patient biopsy tumors from which they were derived. However, it is unclear whether generic Matrigel provides ECM features seen in CRPC-Adeno and CRPC-NEPC tumors. This is important, as tumor type-specific cell-ECM interactions regulate growth and treatment sensitivity in tumor cells. Here, informed by comprehensive transcriptomic and proteomic analysis on patient biopsies, we developed the first synthetic hydrogel-based platform for patient-derived organoid models of CRPC-Adeno and CRPC-NEPC. We investigate the impact of synthetic ECMs on signaling pathways and epigenetic proteins that regulate CRPC to NEPC transformation, as well as their impact on the response of CRPC-NEPC organoids to novel inhibitors that have potential to reprogram and inhibit CRPC-NEPCs. The synthetic hydrogel platform, which presents ECM-specific ligands, can provide a prostate cancer-specific ECM microenvironment and subsequently enable the development of novel single and combinatorial therapeutics.

## Results and Discussion

### CRPC-Adeno patients manifest protein level heterogeneity in cell adhesion proteins

The ECM is a non-cellular component of tissue that provides both biochemical and structural support to its cellular constituents. The ECM is a three-dimensional network of extracellular macromolecules, such as fibrous proteins secreted by resident cells, enzymes, and glycoproteins. Cells adhere to the ECM through the binding of the integrin family of transmembrane receptors that are expressed on cell surfaces^14–16^. Integrins regulate cancer cells’ cellular and molecular fate, including survival, gene expression, and ability to metastasize^17, 18^. Since there are many ECM components as well as integrin receptors, the combination of various expressed ECM components and integrins may, in theory, be quite diverse. In CRPC-Adeno and CRPC-NEPC, the composition and inter-patient diversity of the ECM and integrin components is not well understood. To address this gap, we performed a comprehensive proteomic analysis of cell adhesion proteins using liquid chromatography-tandem mass spectrometry in CRPC-Adeno tumors from 3 patients (primary tumors) and compared to the adjacent non-tumorous prostate tissue (benign). Across all three patients, a higher expression of ECM proteins, including collagen, fibronectin, vitronectin, laminin, and associated integrin subunits α2, α4, α5, and β1 was observed as compared to benign tissue. However, these proteins were upregulated at different levels in each patient, suggesting inter-patient heterogeneity (**Figure 1A**). The proteomics results further indicated an increased protein expression of focal adhesion proteins, such as actin and vimentin, which structurally integrates mechanotransduction networks within cells and is crucial for cancer cell metastasis^19^. These differences motivated us to study a more comprehensive transcriptomic dataset to better understand how differences across CRPC-Adeno and CRPC-NEPC disease state would change the expression of integrin and ECM genes.

**Figure 1.**
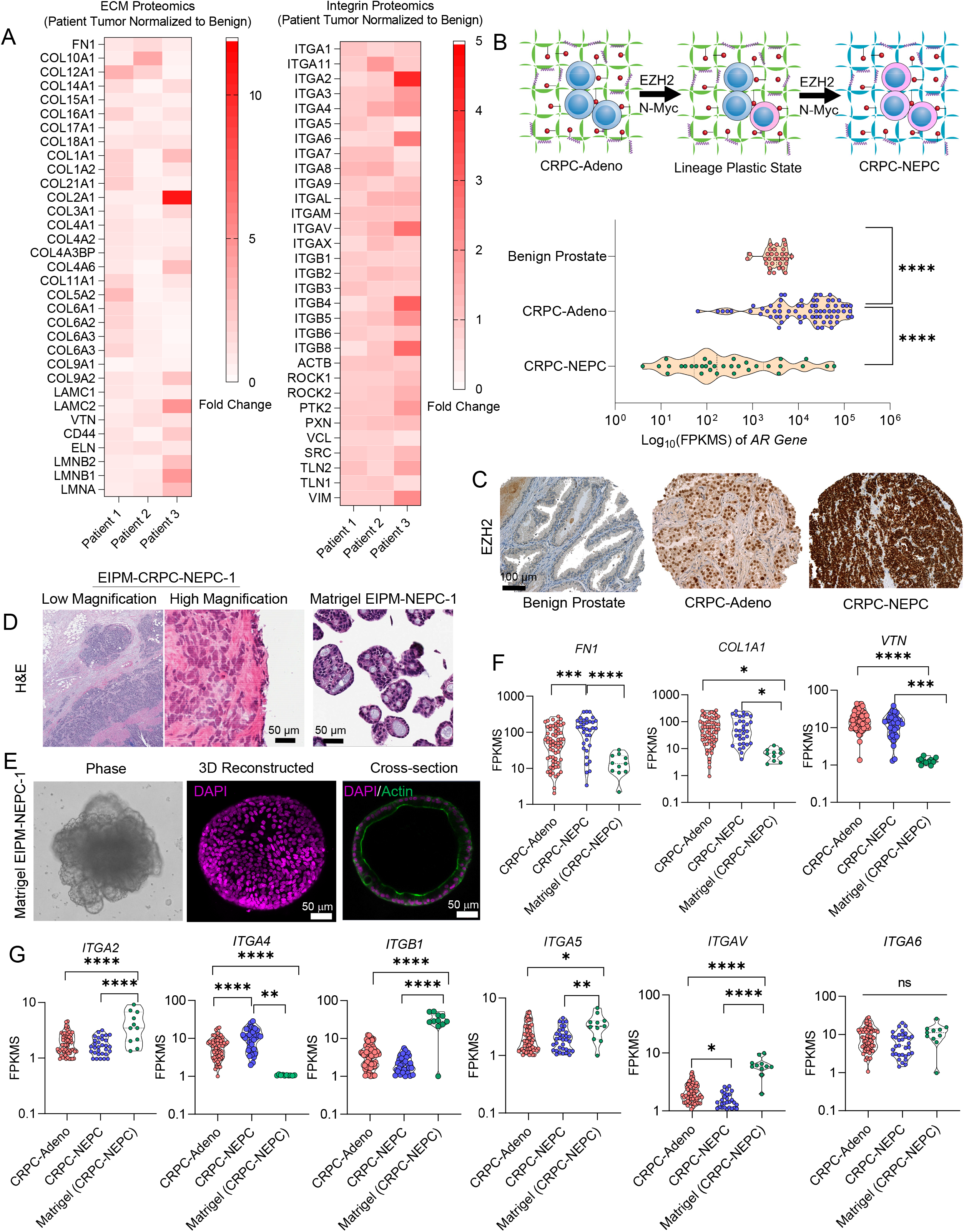
Proteomic and transcriptomic analysis characterizes changes in extracellular matrix and integrins during the progression of prostate cancer. **A)** Mass spectrometry analysis of extracellular matrix (ECM) components (left) and integrin signaling components (right) of primary CRPC-Adeno tumors relative to adjacent normal tissue (n=3, tumor readouts were normalized to adjacent normal tissue). **B)** Schematic of the evolution of CRPC-NEPC from a CRPC-Adeno phenotype (top), which entails the loss of androgen receptor signaling (bottom). **C)** Representative IHC staining demonstrating EZH2 expression in benign prostate, CRPC-Adeno, and CRPC-NEPC patient tumor biopsies. **D)** Representative H&E images of patient EIPM-NEPC-1, metastatic prostate cancer to the liver with overlapping features between small cell neuroendocrine carcinoma and adenocarcinoma: H&E images correspond to frozen tissue submitted for NGS and tissue smear submitted for organoid development, and Matrigel organoids of the same patient tumor (top), **E)** Phase contrast and confocal images of EIPM-CRPC-NEPC-1 organoids cultured in Matrigel. **F)** Transcriptomic expression of ECM components in CRPC-Adeno and CRPC-NEPC patient tumor biopsies, as compared to Matrigel based CRPC-NEPC organoids. **G)** Transcriptomic expression of ECM components in CRPC-Adeno and CRPC-NEPC patient tumor biopsies, as compared to Matrigel-based CRPC-NEPC organoids. (n=74 CRPC-Adeno, n=35 CRPC-NEPC, n=11 Matrigel). For statistical evaluation of all transcriptomic data, groups were compared by a one-way ANOVA, with posthoc Tukey’s test (*p<0.05, **p<0.01, ***p<0.001, and ****p<0.0001).

### Evolution towards a neuroendocrine phenotype drives change in the bioadhesive microenvironment

We studied a dataset of 74 CRPC-Adeno, 35 CRPC-NEPC, and 31 benign tissues from prostate cancer patients. Prior studies suggest that CRPC-NEPCs evolve from CRPC-Adeno (**Figure 1B**) and manifest epithelial plasticity, driven by epigenetic programming of EZH2 and upregulation of N-MYC, resulting in loss of AR signaling dependence^7^. We confirmed the loss of AR signaling dependence by measuring AR expression, observing an expected loss of AR gene expression in CRPC-NEPC (**Figure 1B**). Previously, using whole-exome sequencing (WES) of 114 metastatic tumor-normal tissue pairs, we have demonstrated that the mutational landscape of CRPC-NEPC tumors was similar to that of CRPC-Adeno. However, it also included frequent loss of *RB1*, which encodes the retinoblastoma tumor suppressor protein (in 70% of CRPC-NEPC versus 32% of CRPC-Adeno samples), and mutation or deletion of *TP53*, which encodes the p53 tumor suppressor protein (in 66.7% of CRPC-NEPC versus 31.4% of CRPC-Adeno samples)^7^. Importantly, the transdifferentiation from CRPC-Adeno to CRPC-NEPC is strongly correlated with Polycomb group protein-mediated epigenetic silencing, largely owing to the upregulation of epigenetic modifier EZH2, which methylates histone H3 lysine 27 (H3K27) to alter the expression of lineage specification genes, cell cycle checkpoint genes, and DNA repair genes^1, 7^ Immunohistochemistry analysis of EZH2 in patient biopsies confirmed that EZH2 protein was more abundant in CRPC-NEPC than in CRPC-Adeno, and absent in benign prostate tissue (**Figure 1C**).

To better understand the expression of ECM and integrins in CRPC-Adeno and CRPC-NEPC patients, we interrogated RNA sequencing results from 109 patient tumors, including 74 patient tumors with clinical and histologic features of CRPC-Adeno and 35 with features of CRPC-NEPC, as confirmed by pathologic consensus criteria^8^. We also analyzed RNA sequencing from 11 independent CRPC-NEPC Matrigel organoids derived from two CRPC-NEPC patients, reported earlier by our group as OWCM154 and OWCM155^13^. In that study, fresh tumor tissues from 25 patients with metastatic prostate cancer were used for Matrigel-based organoid development with an overall patient success rate of 16% (4/25)^13^. In one of these cases, referred to as EIPM-CRPC-NEPC-1 (OWCM155), Matrigel organoid was successfully developed from fresh tissue obtained from a metastatic biopsy from a patient with prostatic adenocarcinoma Grade Group 5 (Gleason Score 4+5=9). In the metastatic biopsy to the liver, overlapping features between small cell carcinoma and adenocarcinoma were seen (**Figure 1D**). The Matrigel-derived EIPM-CRPC-NEPC-1 demonstrated both similar cytomorphology and gland formation (**Figure 1D**). Confocal microscopy indicated the luminal gland-like morphology of Matrigel-derived EIPM-CRPC-NEPC-1 organoids (**Figure 1E**). Next, comparative analysis of RNA sequencing readouts from CRPC-Adeno and CRPC-NEPC patients revealed an upregulated expression of ECM genes in CRPC-NEPCs, including fibronectin *(FN1)*, in contrast to Matrigel organoids of CRPC-NEPCs (**Figure 1F**). Nevertheless, the expression levels of ECM genes in both CRPC-Adeno and CRPC-NEPC patients showed a wide distribution, suggesting that an ideal organoid should match the expression of ECM in a patient-specific manner (**Figure 1F**). We observed notable differences in integrin expression between Matrigel-derived CRPC-NEPC organoids and patient CRPC-NEPC and CRPC-Adeno samples. Specifically, genes encoding for integrin α2, β1, and αV were upregulated in Matrigel organoids relative to the patient cohort, whereas integrin α4 was not expressed in Matrigel **(Figure 1G, Supplementary Fig. 1)**. The collective proteomic and RNA sequencing highlights (a) the heterogeneity of expression of different ECM proteins and integrin ligands that the cells bind to in prostate cancer patients, suggesting potential putative ECM cues that guide the growth of prostate tumors and are needed in a prostate cancer organoid (b) the limited ability of CRPC-NEPC Matrigel-derived organoids to recapitulate ECM expression seen in CRPC-NEPC patients.

### CRPC-Adeno Matrigel-derived organoids from patient tumor biopsy capture *in situ* histological features and hallmark gene expression

To further compare CRPC-NEPC organoids with CRPC-Adeno, we first derived a new CRPC-Adeno Matrigel-derived organoid from a fresh patient biopsy. The primary tumor tissue **(Figure 2A)** was obtained from a rapid autopsy^20^, allowing for the establishment of multiple organoid lines from various tumor sites. CRPC-Adeno Matrigel-derived organoids were derived as demonstrated earlier by our group for CRPC-NEPCs^13^, and after 5 subsequent passages, we confirmed the identity of the CRPC-Adeno tumors in organoids by comparing gene expression, using NanoString, against known molecular markers of CRPC-Adeno **(Figure 2B)**. Among these organoids, only the lymph node biopsy site expressed the markers associated with CRPC-Adeno without developing into a neuroendocrine phenotype. The tumor organoid, referred to as EIPM-CRPC-Adeno-1 was verified as being CRPC-Adeno by multiple molecular markers **(Figure 2C)**. Next, immunohistochemistry analysis indicated that both patient’s tumor sample and tumor organoid manifested comparable histological features (Hematoxylin and eosin stain) as well as surface marker immune profile (AR, PSMA, and NKX3.1) **(Figure 2C)**. When EIPM-CRPC-Adeno-1 organoids were compared with established CRPC-NEPC organoids in Matrigel (EIPM-CRPC-NEPC-1 and EIPM-CRPC-NEPC-2) **(Figure 2D)**, the CRPC-Adeno organoid showed strikingly distinct genes than neuroendocrine derivative, for example maintaining expression of *AR, KLK3, ENO2, NKX3, AR-V1, and FOLH1 (PSMA)*.

**Figure 2.**
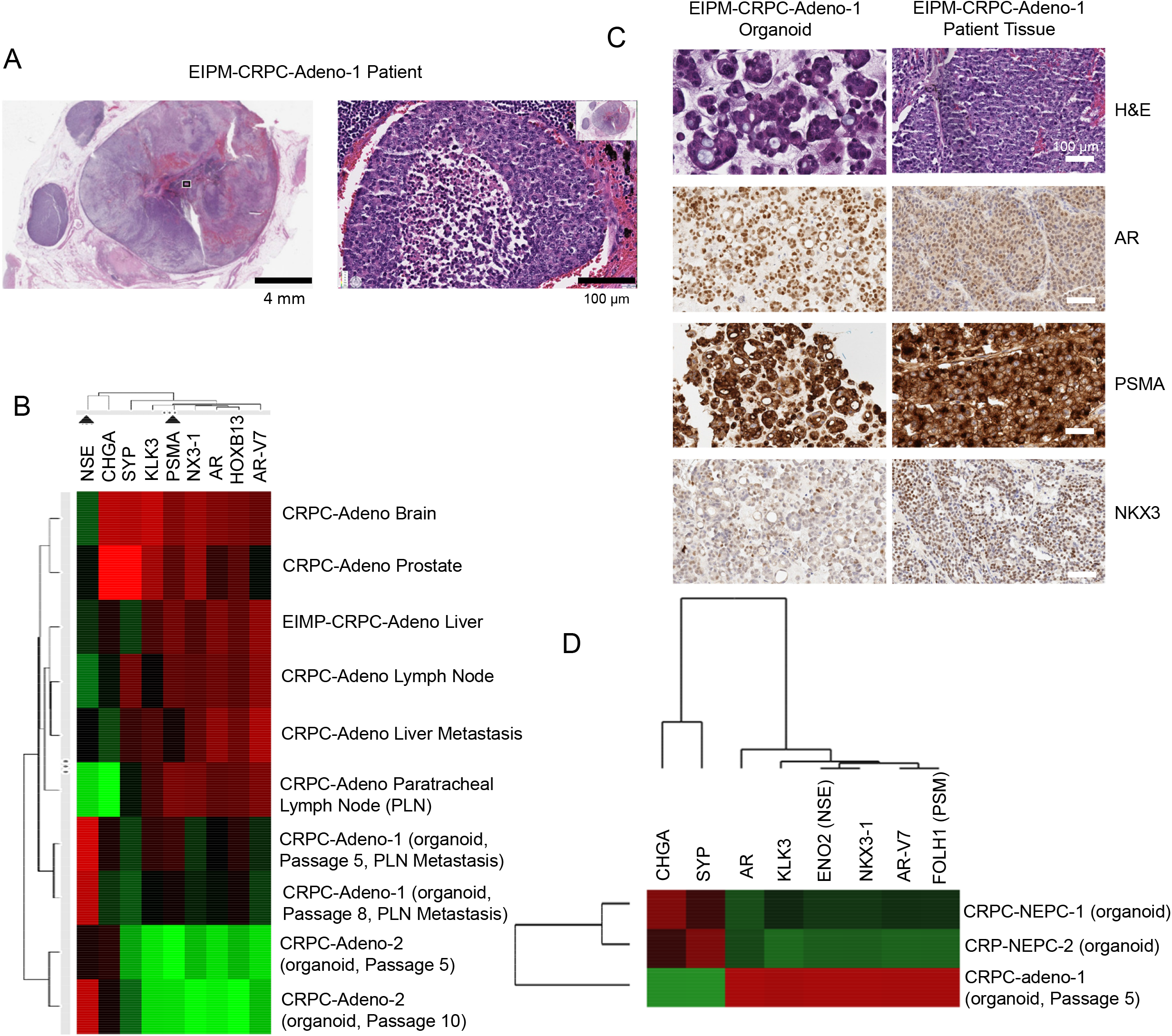
Development and characterization of a new CRPC-Adeno organoid from a patient tumor biopsy. **A)** H&E staining of a rapid autopsy specimen from a single EIPM-CRPC-Adeno patient. **B)** Nanostring analysis of hallmark gene expression in multiple metastatic sites and derived Matrigel organoids. **C)** H&E and IHC staining of CRPC-Adeno patient and Matrigel organoid that demonstrated CRPC-Adeno gene signature. **D)** Nanostring-based comparative analysis of prostate cancer-associated genes among CRPC-Adeno and CRPC-NEPC Matrigel organoids.

### Bioengineered synthetic hydrogel-based organoids of CRPC-Adeno and CRPC-NEPC promote an invasive phenotype

The RNA sequencing analysis of ECM and integrins in prostate cancer patients showed that integrins and ECM genes were differentially expressed in CRPC-NEPC organoids relative to their patient tumors. This could be attributed to the challenges presented by Matrigel organoids as they are not conducive to controlled modifications^21, 22^ to present ligands of interest, and are embedded in a protein-rich ECM that confounds the understanding of the role of an individual or combinatorial ECMs and corresponding integrins found in patients. This motivated us to engineer a synthetic hydrogel-based organoid platform to grow prostate cancer and more faithfully recapitulate the interactions between the CRPC-Adeno and CRPC-NEPC with their respective ECMs. To engineer a platform technology that responds to CRPC-Adeno and CRPC-NEPC ECM needs, we further examined the metalloproteinases (MMPs) secreted by prostate cancer cells. MMPs promote cancer progression through metastasis, proliferation, apoptosis, angiogenesis, and EMT^23^. Using RNA sequencing, we examined the expression levels of a wide range of MMPs and observed that MMP-1, 9, 13, 15, and 16 were highly expressed in CRPC-NEPC as compared to benign prostate tissue or CRPC-Adeno tumors (**Figure 3A**), whereas MMP-2, 3, 7, 14, 17, 19, 20, and 26 were either downregulated or unchanged **(Supplementary Fig. 2)**. MMP-9 and −13 were further upregulated in CRPC-Adeno patient tissues, with MMP-9 comparable to CRPC-NEPC levels. In contrast, MMP levels in primary, localized prostate cancer samples (PCa) were often similar to benign prostate tissues (**Figure 3A, Supplementary Fig.2)**.

**Figure 3.**
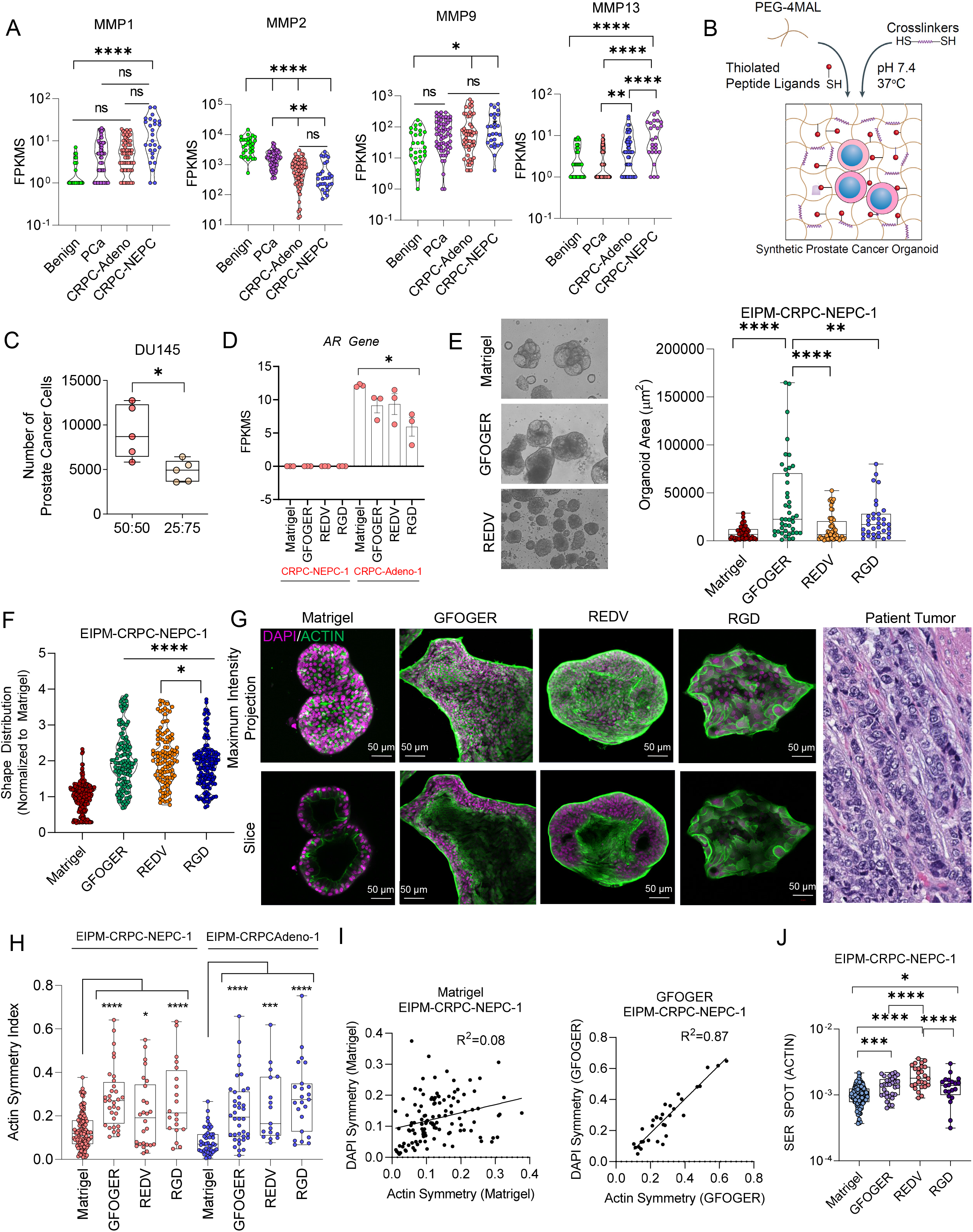
Development and characterization of CRPC-NEPC PEG-4MAL synthetic organoids from patient-derived tumors. **A)** Transcriptomic analysis of the patient cohort for matrix metalloproteinases. **B)** Schematic of prostate tumor tissue expanding and cleaving the tumor niche (n=31 BenignProstate, n=66 primary, localized prostate cancer (PCa), n=74 CRPC-Adeno, n=35 CRPC-NEPC, n=11 Matrigel). **C)** Flow cytometry analysis of DU145 prostate cells in organoids grown under tunable degradability (n=5 per condition). **D)** Androgen receptor expression of CRPC-Adeno and CRPC-NEPC organoids (left) (n=3 per condition). **E)** Representative phase-contrast imaging of organoids in Matrigel and our PEG-4MAL platform (middle), and measurement of the organoid area across conditions. **F)** Representative high content imaging quantification of prostate organoid shape distribution across ECM conditions (Matrigel n=204, RGD n=135, REDV n=113, GFOGER n=123). **G)** Left, representative confocal imaging of organoid morphology across ECM conditions, with DAPI in purple and actin in green. Right, comparison of morphology from high-content imaging studies, compared to CRPC-NEPC tissue from a patient. **H)** DAPI symmetry across organoid culture conditions from high content imaging (CRPC-NEPC: Matrigel n=128, GFOGER n=32, REDV n=26, RGD n=20. CRPC-Adeno: Matrigel n=45, GFOGER n=38, REDV n=17, RGD n=23). **I)** Correlation between DAPI and Actin symmetry between Matrigel and GFOGER organoids (Matrigel n=128, GFOGER n=32, REDV n=26, RGD n=20). **J)** Texture analysis of actin morphology among organoids. For all comparisons, groups were compared by a one-way ANOVA, with posthoc Tukey’s test. For *p<0.05, **p<0.01, ***p<0.001, and ****p<0.0001.

We, therefore, chose to engineer a cell-laden hydrogel-based organoid where the ECM ligand functionalized polymers can be crosslinked into a 3D scaffold using network crosslinkers that were MMP-9 and 13 cleavable peptide sequences. We exploited the chemistry between a 4-arm Maleimide-functionalized Polyethylene glycol (PEG-4MAL) that clicks with thiol moieties on other crosslinking materials, such as ECM peptide mimics and MMP-degradable peptides, using a click chemistry^22, 24^ Out of the four Maleimide arms of PEG-4MAL, one-arm can be functionalized with ECM-mimicking peptide ligand of interest using the maleimide-thiol chemistry (**Figure 3B**). The remaining three arms of PEG-4MAL can be crosslinked with dithiolated MMP-9- and MMP-13-degradable peptides (GCRD**VPM**↓SMRGGDRCG, referred as VPM hereafter). However, a hydrogel only crosslinked with MMP-degradable peptide can quickly biodegrade due to secreted proteases, and therefore we further optimized the stability of hydrogels by mixing VPM with non-degradable crosslinker dithiothreitol (DTT). To determine the effect of MMP-mediated degradability of organoid on tumor growth, we tested a model prostate cancer cell line (DU145), which is AR-negative, for growth under PEG-4MAL matrix crosslinked with a mixture of 50% VPM and 50% non-degradable DTT, or 25:75 ratio of VPM:DTT. These hydrogels were functionalized with a fibronectin-mimicking tri-amino acid sequence, arginine-glycine-aspartate, or RGD because Matrigel includes naturally occurring RGD adhesion domains at ~0.7 μM concentration^25^. We chose RGD peptide at 3 mM concentration to maximize adhesion sites for prostate cancer cell binding. A 50:50 ratio of VPM to DTT supported the growth of prostate cancer cells over 7 days without significantly degrading the gels, whereas 25% VPM crosslinked hydrogels had limited growth (**Figure 3C**). VPM and DTT have different molecular weights and sizes, and therefore the ratio could potentially impact hydrogel porosity, which can impact regular cellular processes such as growth and spread. A 100% VPM led to the rapid degradation of hydrogels over 48 hours and was not tested further. AR-dependent cell line LNCaP responded similarly to the organoid degradation effect (**Supplementary Fig. 3A**). Therefore, all further studies considered 50% VPM to accommodate optimal growth conditions for CRPC-NEPCs and CRPC-Adeno derived organoids, as well as maintain stable hydrogel organoids.

Next, we determined the effect of three individual ECM components (fibronectin/vitronectin mimicking RGD, fibronectin mimicking REDV, and collagen mimicking GFOGER peptide), a mix of all three ECMs, and Matrigel on the growth of EIPM-CRPC-NEPC-1 tumor cells. Here, the CRPC-NEPC patient cells were derived first in Matrigel, serially passaged and validated for hallmark signature of CRPC-NEPC tumors, and then serially implanted into PEG-4MAL hydrogels. The proteomics analysis of patient tumors (**Figure 1**) indicated the upregulation of collagen, fibronectin, and vitronectin, among other ECM proteins. We functionalized PEG-4MAL macromers with GFOGER, a triple helical synthetic peptide derived from type I collagen with high binding affinity for α1β1, α2β1, α10β1 and α11β1^26, 27^, REDV, a tetrapeptide Arg-Glu-Asp-Val that mimics Fibronectin in its ability to bind α4β1 integrins, and RGD is a short linear peptide present in vitronectin, fibronectin and other ECM proteins that bind several integrins, including αvβ3, αvβ1, and α5β1^28-30^. After seven days of culture in PEG-4MAL, Ki-67 analysis indicates similar proliferative behavior of CRPC-NEPC tumors across collagen mimetic (GFOGER), fibronectin mimetic (REDV), and fibronectin/vitronectin mimetic (RGD), Matrigel, as well as a combination of GFOGER (75%) with RGD (10%) and REDV (15%) that mimicked the ratios from patient proteomics studies (**Figure 1A**). All matrices demonstrated similar proliferative potential, with a modest increase in the combination ECM **(Supplementary Fig. 3B)**. These results verify that the synthetic hydrogels provide a niche that allows for cell proliferation in a manner consistent with Matrigel, however, unlike Matrigel PEG-4MAL hydrogels are more defined in their composition.

A key distinction between CRPC-Adeno and CRPC-NEPCs is the expression of *AR* gene: CRPC-Adeno express AR but lose AR expression upon transition to CRPC-NEPCs. Accordingly, mRNA analysis indicated that AR expression was retained after the growth of CRPC-Adeno in all four ECM conditions (Matrigel, REDV, GFOGER, and RGD), whereas AR expression was absent in CRPC-NEPCs among all conditions, verifying that these organoid matrices maintain the disease phenotype (**Figure 3D**). Additionally, we observed that the ECM type influenced the organoid growth area of CRPC-NEPCs, defined as the total 3D surface area of a cell cluster as measured by confocal imaging. GFOGER led to a maximal organoid area together with a variable area size distribution compared to Matrigel. We hypothesize that ECM ligands will upregulate gene expression programs associated with growth signaling, resulting in increased cluster area and changes in morphology. In contrast, REDV and RGD functionalized matrices led to a significantly smaller organoid area than GFOGER and remained similar to Matrigel (**Figure 3E**).

In CRPC-NEPC patients, we had observed irregularity in tumor organization (**Figure 1D**), which is characteristic of small cell carcinomas and occurs as prostate tumors lose luminal morphology and transform to irregular shapes as compared to CRPC-Adeno^31^. However, the Matrigel organoids induced luminal morphology in CRPC-NEPCs (**Figure 1E**). We hypothesized that the ECM type may regulate the non-uniformity in cancer cell clusters within each hydrogel. High-content imaging using Operetta™ and cell cluster or shape analysis confirmed that Matrigel induced uniformly spherical organoids. The same imaging analysis revealed that GFOGER, REDV, and RGD matrices led to a wide range of non-spherical shapes of tumor clusters formed with both EIPM-CRPC-NEPC-1 and EIPM-CRPC-Adeno-1 patient-derived cells (**Figure 3F, Supplementary Fig. 4**). The shape of organoids was evaluated by considering sphericity (shape) of the organoids, which is a measure of how closely the organoids 3D volume approximates a sphere. When normalized to Matrigel, all three matrices showed nearly a 2-fold change in shape, indicating an increase in the overall sphericity of the synthetic organoids and changing the underlying morphology. These observations were further validated using confocal imaging, which confirmed that tumors display luminal morphology in Matrigel CRPC-NEPC organoids (**Figure 3G**), where the other three matrices induced distinct, invasive-like morphology and reflected EMT transition. In GFOGER and RGD organoids, we observed increased protrusion of cell and its cytoskeletal protein, actin, along with a less luminal phenotype. This irregularity in shape is reflective of patient tumors (**Figure 3G**).

Remodeling of the actin cytoskeleton is necessary for EMT; however, to understand how this is regulated in organoids functionalized with CRPC-NEPC- and CRPC-Adeno-specific ECM ligands, we performed image analysis on organoids stained with actin and DAPI, using the PhenoLOGIC™ machine-learning tool in Operetta high-content imaging system. The machine-learning tool quantifies texture parameters (Spots or Ridges) based on the occurrence of a characteristic intensity pattern within the image whilst the symmetry morphology parameter quantifies the distribution of either texture or fluorescence intensities inside a region of interest. The cellular texture parameters can reflect the cytoskeletal status of the cells^32^. Compared to Matrigel, EIPM-CRPC-NEPC-1 tumors grown in all three ECMs demonstrated a broader fluorescence distribution concerning cellular symmetry of actin filaments (**Figure 3H**). We examined the correlation between actin symmetry of CRPC-NEPCs (based on actin staining) and nuclear symmetry and observed high correlation between the two parameters in GFOGER (R^2^=0.87), REDV (R^2^=0.84), and RGD (R^2^=0.93) but the poor correlation in Matrigel organoids (R^2^=0.08) (**Figure 3I, Supplementary Figure 5B**). In addition to cell morphology features, texture parameters of the cytoskeletal fibers at the 1px scale were determined to measure the differences in cytoskeletal and nuclear structure. Once again, the synthetic hydrogel-based organoids with tumor-specific ECM resulted in significantly higher actin spots and ridges in CRPC-NEPC tumors compared to Matrigel (**Figure 3J, Supplementary Figure 6**). However, in contrast to the pattern seen with actin symmetry, CRPC-Adeno grown in synthetic ECMs did not show an increased texture compared to Matrigel (Supplementary Fig. 6A, B). We observed that these organoids generated a wider variety of nuclear and extracellular morphologies and the differences appear to be mediated by ECM type. Collectively, these studies suggest that the actin cytoskeleton is dynamically reorganized in synthetic ECMs and allude to the possibility that in synthetic ECMs, tumor cells acquire increased EMT signature, cell-matrix interactions, and enhanced migratory and invasive capabilities. Since these features are seen in patient tumors, synthetic ECMs may be able to provide more accurate models than widely used Matrigel.

### PEG-4MAL organoids modulate the expression of the epigenetic regulator EZH2 and response to epigenetic therapies

Progression in prostate cancer, specifically from CRPC-Adeno to CRPC-NEPC, is associated with changes of epigenetic regulators. In patients, the expression of EZH2 increases with the neuroendocrine disease as compared to CRPC-Adeno and prostate adenocarcinoma (**Figure 4A**). When grown in Matrigel, CRPC-NEPC organoids retain high expression of EZH2 expression, as validated using immunohistochemistry (**Figure 4B**). We observed that while EZH2 expression was spatially heterogenous in the patient sample, it was consistent and uniform in Matrigel-based CRPC-NEPC organoids from the same patient. Previous studies have shown that matrix properties, including stiffness, can modulate nuclear accessibility and epigenetics^33^. We wondered whether ECM-integrin interactions may regulate the EZH2 expression and its activity in methylating histone. This hypothesis was driven by our observation that synthetic organoids generate a more spread phenotype (**Figure 3**), indicating that progression (and consequently epigenetic regulation) are governed in part by extracellular matrix presentation. Both GFOGER and especially REDV peptide presentation significantly increased the expression of EZH2 and its target histone modification H3k27Me3, as compared to Matrigel (**Figure 4C**). In contrast, RGD and the combination ECM had no significant increase over Matrigel (**Figure 4D**). The overall and consistent increase in expression of both EZH2 and H3K27Me3 led us to conclude that epigenetic regulators were being modulated by interactions of prostate cancer cells with the ECM ligands.

**Figure 4.**
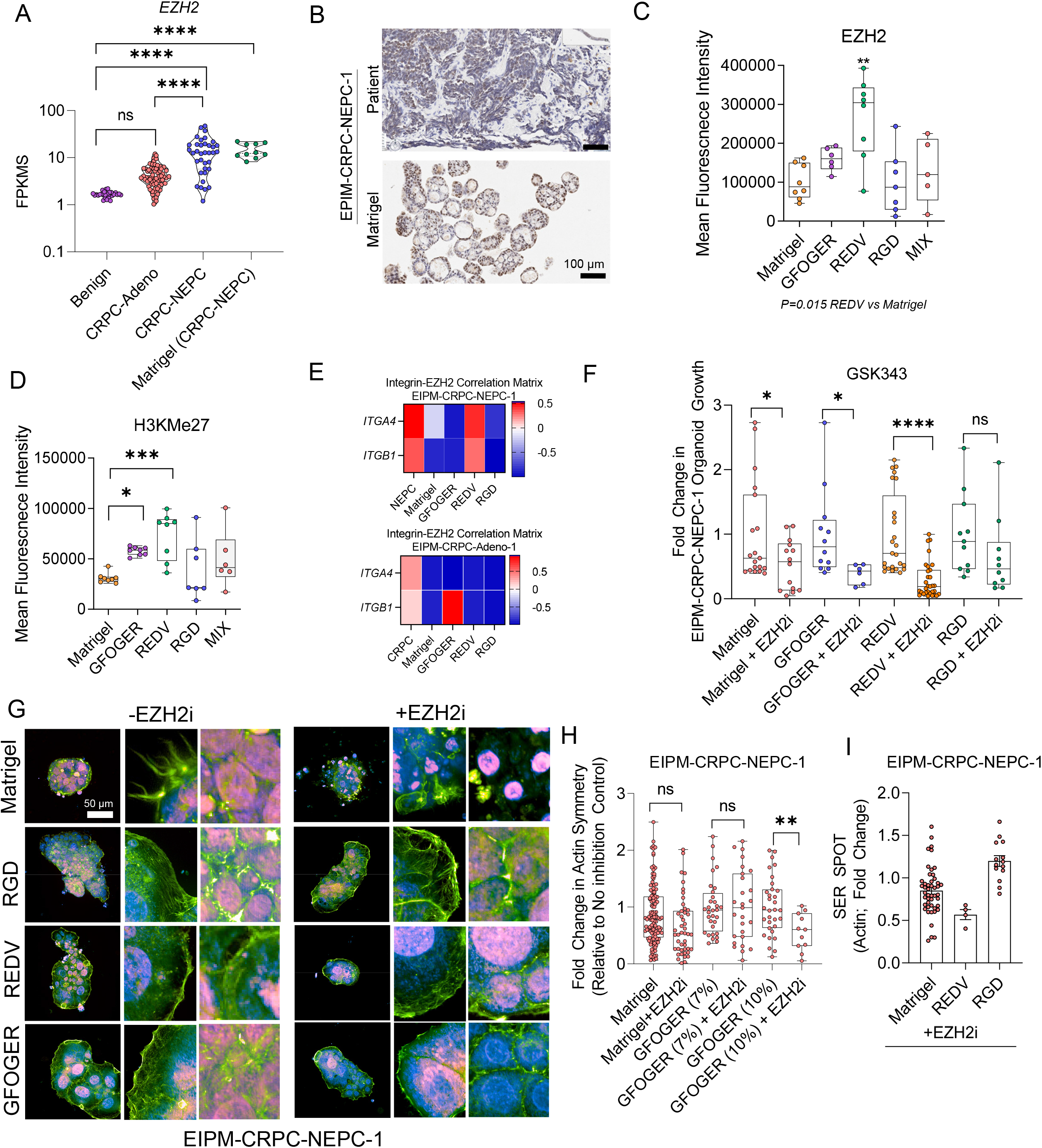
Epigenetic regulator EZH2 is governed by the extracellular matrix microenvironment. **A)** Transcriptomic analysis of EZH2 across patient subtypes during disease progression (Benign Prostate n=29, CRPC-Adeno n=66, CRPC-NEPC n=36, Matrigel n=10). **B)** Representative IHC and immunofluorescence for patient and organoid samples. **C)** Flow cytometry analysis of EZH2 across organoid conditions (n=5 per condition). **D)** Flow cytometry analysis of H3K expression across organoid conditions (n=5 per condition). **E)** Integrin and EZH2 correlation matrices for neuroendocrine (top) and castration-resistant (bottom) organoids. **F)** Metabolic base measurement of organoid viability under treatment with an EZH2 inhibitor, GSK343 (Matrigel n=19, Matrigel+EZH2i n=14, GFOGER n=12, GFOGER+EZH2i n=6, REDV n=25, REDV+EZH2i n=31, RGD n=11, RGD+EZH2i n=10). Treated and untreated comparisons were evaluated by a two-tailed *t*-test with *p<0.05 and ****p<0.0001. **G)** Representative images of organoids under confocal imaging with and without EZH2 inhibition. **H)** Quantification of actin symmetry from high content imaging (Matrigel n=128, Matrigel+EZH2i n=49, GFOGER 7% n=32, GFOGER 7%+EZH2i n=27, GFOGER 10%+EZH2i n=32, GFOGER 10%+EZH2i=11). **I)** Actin texture analysis of CRPC-NEPC organoids treated with EZH2 inhibitor (Matrigel n=49, REDV n=4, RGD n=14). For all comparisons, unless otherwise noted, a one-way ANOVA, with posthoc Tukey’s test was performed with *p<0.05, **p<0.01. **G)** Texture analysis of actin morphology changes due to EZH2 inhibition among CRPC-NEPC organoids. For all comparisons, a one-way ANOVA, with posthoc Tukey’s test was performed with *p<0.05.

Having uncovered these interactions at the cellular level, we next sought to understand the interplay between EZH2 and the ECMs at the transcriptomic level. To test our hypothesis that ECM-integrin interactions would modulate the expression of EZH2, we performed correlation analysis on mRNA levels between integrin subunits and EZH2. This analysis entailed determining the correlation coefficient between EZH2 and integrin associated genes from whole transcriptome sequencing analysis and comparing these trends from patients to organoids. The analysis allows for a determination of whether gene expression correlation is maintained in organoids, as compared to patients stratified by disease type.

There was a positive correlation between α4 and β1 integrins in REDV matrices and the patient samples, but not in other matrices **(Figure 4E)**. In contrast, integrins and EZH2 in EIPM-CRPC-Adeno-1 organoids showed a low correlation score, and therefore we continued with only CRPC-NEPC for EZH2 inhibition studies.

We have previously shown that the treatment of CRPC-NEPCs with the EZH2 inhibitor GSK343 significantly decreases the growth of CRPC-NEPC tumors in Matrigel organoids^13^. To further test our hypothesis that EZH2 inhibition would be dependent on ECM ligand types, we treated our organoids with GSK343 and evaluated growth using a luminescence assay **(Figure 4F)**. When compared to untreated controls, all organoids except RGD responded to EZH2 inhibition with diminished growth. Correlating with high EZH2 expression, treatment with EZH2 inhibitor resulted in a significantly greater reduction in proliferation in organoids functionalized with REDV ligands, highlighting an increased sensitivity to the drug. These results corroborate flow cytometry results that may allude to the potential role fibronectin (REDV ligand) could play in increasing the dependence of CRPC-NEPCs on EZH2.

Next, we characterized the role of EZH2 inhibition on organoid protein localization using confocal microscopy (**Figure 4G**). In comparison to untreated organoids, CRPC-NEPCs in Matrigel underwent a loss of actin cytoskeleton structure when treated with an EZH2 inhibitor, whereas synthetic organoids did not show the same phenotype. Notably, Matrigel organoids had uniform EZH2 expression throughout the nucleus. Contrary to that, the PEG-4MAL organoids showed heterogeneous expression of EZH2 across the nucleus, with differences being especially abundant among REDV and GFOGER organoids.

To quantify morphological differences due to EZH2 inhibition, we performed high content imaging of organoids under EZH2 inhibition **(Figure 4H)** and did not observe a significant difference in actin symmetry with inhibition of EZH2. We further tested whether organoid stiffness could also play a role in regulating EZH2 inhibitor response because tissue stiffness could modulate cytoskeletal elements of cells. Intriguingly, we observed that changes in the ECM stiffness (by increasing PEG-4MAL weight %) resulted in a significant reduction in actin symmetry upon EZH2 inhibition. These results highlight a potential interaction between stiffness and EZH2 and warrant further investigation. Furthermore, we performed a texture analysis to reflect on the cytoskeletal status of the cells^32^ and uncovered that REDV organoids had a reduction of the actin symmetry under EZH2 inhibition, demonstrating that morphological differences are dependent on underlying ECM-integrin ligations. These results validate that, morphologically, synthetic ECM-grown organoids express both greater actin symmetry as well as capture a broader spectrum of cluster morphologies.

### The ECM composition of PEG-4MAL organoids regulates the transcriptional signature of CRPC-NEPCs

To further explore the effect of ECM components in synthetic ECM organoids on transcriptional programming, we performed bulk RNA sequencing on PEG-4MAL and Matrigel-grown CRPC-Adeno and CRPC-NEPC organoids. Each PEG-4MAL ECM condition was normalized to Matrigel for comparison with Matrigel and across synthetic organoids. Unsupervised analyses were performed to determine whether synthetic ECM organoid models express common genes or potential regulatory mechanisms and found that the majority of genes differentially expressed between an ECM and Matrigel clustered together in GFOGER and REDV but were distinct from RGD organoids (**Figure 5A**). This initial observation hinted at potential reasons why the EZH2 inhibitor response was not significantly different from untreated controls in RGD organoids (**Figure 4F**). Using - log2fc>1 and padj<.05 or log2fc <-1 and padj<.05 as thresholds, we found that 62 shared genes were upregulated in EIPM-CRPC-NEPC-1 tumors grown in all three ECM relative to Matrigel (**Figure 5B**). These genes were expressed to a similar extent in GFOGER and REDV functionalized organoids and even higher levels in RGD functionalized organoids (**Figure 5B)**. In contrast, EIPM-CRPC-Adeno-1 showed upregulation of only 6 differentially expressed genes (**Figure 5C)**. RNA sequencing further revealed that RGD organoids induced changes to 84 uniquely expressed genes in EIPM-CRPC-NEPC-1 tumors, of which 77 were upregulated and 7 downregulated (**Figure 5D**). These genes were unique to tumors grown in RGD matrices only and were not found to be different in GFOGER and REDV hydrogels. The GFOGER organoids, on the other hand, upregulated 11 unique genes and downregulated 15 genes (**Figure 5E**). The REDV organoids had even fewer changed genes than Matrigel (**Supplementary Figure 7**), with 12 uniquely upregulated genes and 4 downregulated genes. In contrast, the gene mobilization in CRPC-Adeno was exceptionally higher than the gene mobilization seen in CRPC-NEPCs in RGD (379 CRPC-Adeno, 84 CRPC-NEPC) (**not shown**). CRPC-Adeno grown in GFOGER also showed an upregulation of 160 genes and downregulation of 99 genes (**Supplementary Fig. 8**), whereas REDV induced 30 unique upregulated genes and 11 downregulated genes (**Supplementary Figure 7**). These results highlight that different types of ECM mobilize distinct genes in CRPC-Adeno and CRPC-NEPC, which could correlate with differences in therapeutic drug response for many drugs.

**Figure 5.**
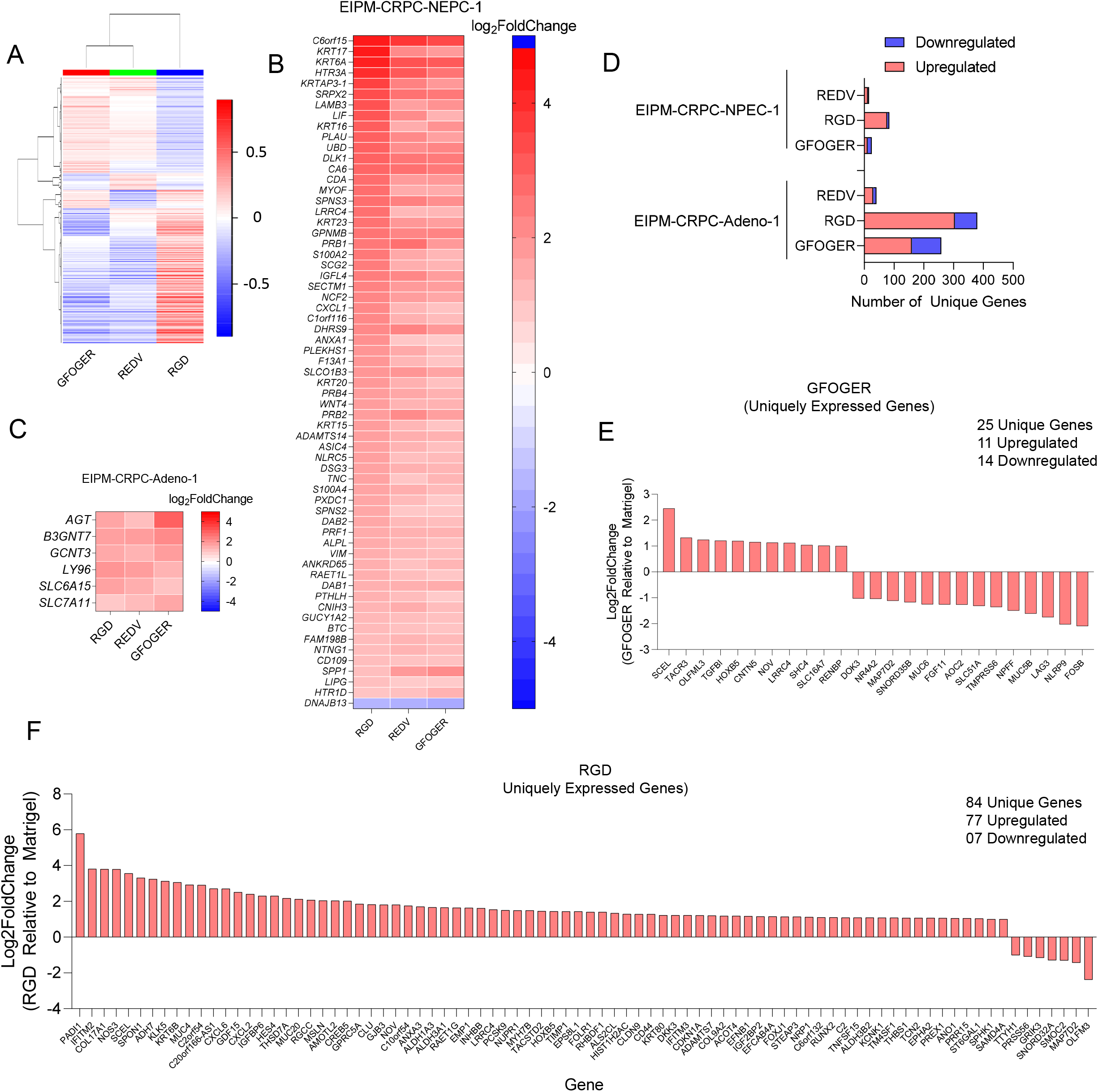
Differential gene expression unveils that microenvironment enhances differing profiles for prostate cancer. **A)** Heatmap of differentially expressed genes in prostate cancer across synthetic hydrogel-based organoid niches. **B)** Differentially expressed genes expressed by synthetic organoids in CRPC-NEPC, relative to Matrigel. **C)** Differentially expressed genes expressed by synthetic organoids in CRPC-Adeno, relative to Matrigel. **D)** Quantification of differentially expressed genes in synthetic organoids relative to Matrigel. **E)** Single unique genes expressed by GFOGER organoids in CRPC-NEPC. **F)** Single unique genes expressed by RGD organoids in CRPC-NEPC. All groups were analyzed from the whole transcriptome sequencing of n=3 organoids per condition.

### ECM differentially regulates gene expression pathways in prostate cancer organoids

Next, we performed gene set enrichment analysis (GSEA) to assess gene expression pathways associated with each synthetic ECM (**Figure 6, Supplementary Figure 9**). The enriched pathways and hallmarks were identified by pre-ranked GSEA using the gene list ranked by log-transformed P values with signs set to positive/negative for a fold change of >1 or <1, respectively. GSEA on multiple pathways associated with CRPC-NEPCs revealed that PEG-4MAL organoids up-regulated extracellular receptor, EMT transition, epigenetic regulator, and EZH2 target pathways as compared to Matrigel organoids (**Figure 6A**). These pathways had a false discovery rate (FDR) q-value less than 0.25, which is a significant measure in the analysis of genome-wide expression data. The CRPC-Adeno organoids, on the other hand, enriched for EMT in all three matrices and epigenetic pathways were only enriched in REDV and GFOGER. ECM genes were not enriched in REDV and GFOGER and were significantly enriched in RGD relative to Matrigel.

**Figure 6.**
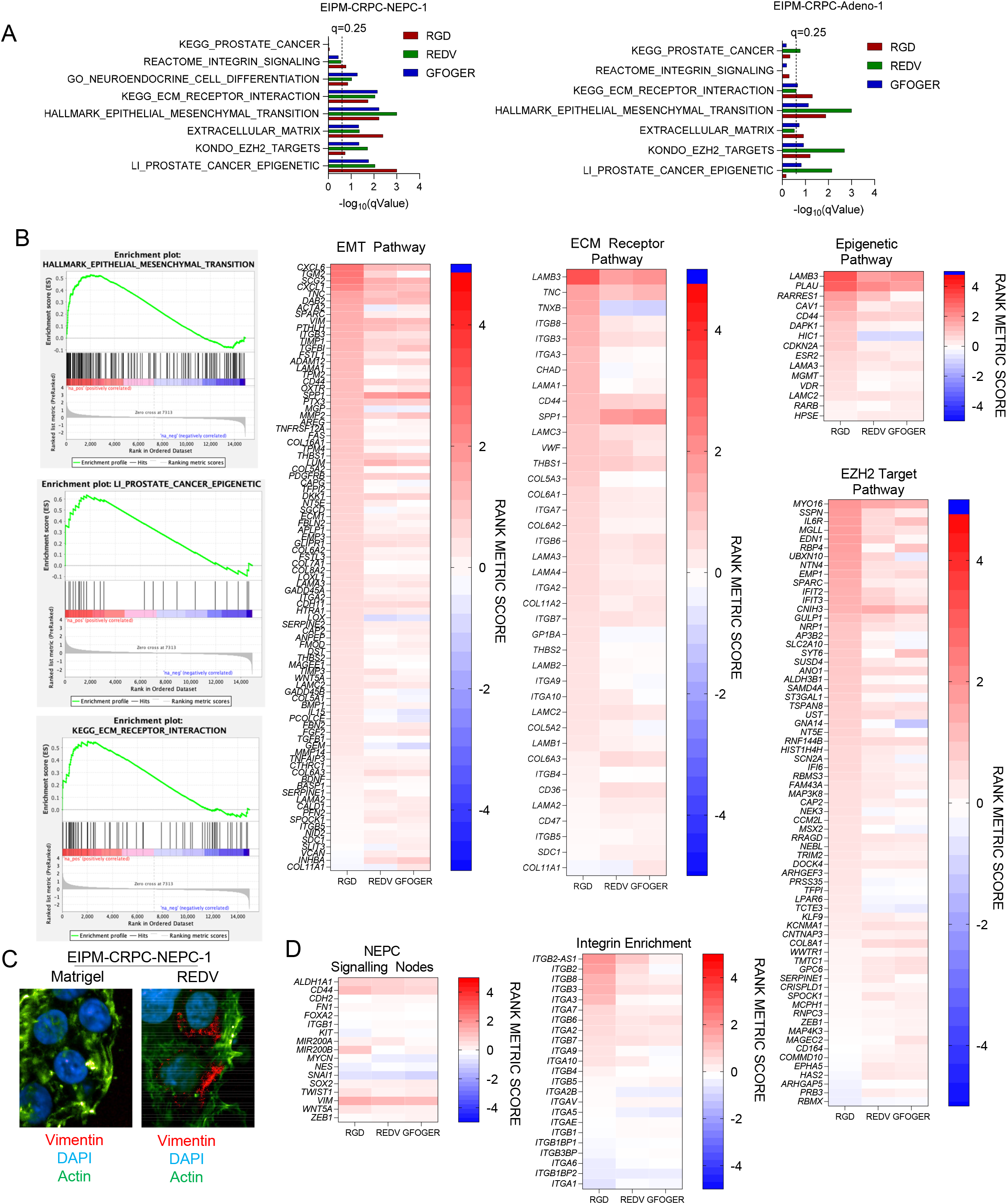
Extracellular matrices in synthetic hydrogels drive tumors towards a transcriptionally distinct phenotype. **A)** q-Values of four GSEA pathways for (left to right) epithelial to mesenchymal transition, prostate cancer epigenetic markers, targets of the epigenetic regulator EZH2, and ECM interaction with all comparisons to Matrigel. A q-value below 0.25 is considered significant. **B)** Left, cumulative enrichment score of organoids. Right, heat maps of enriched genes for the above, with comparisons performed relative to Matrigel. From left to right: the epithelial to mesenchymal transition pathway, the ECM receptor pathway, the epigenetic pathway, and targets of the epigenetic regulator EZH2. **C)** Representative confocal imaging of organoids for vimentin (red, an EMT marker), DAPI (blue), and actin (green). **D)** Enrichment of signaling nodes for neuroendocrine prostate cancer. All sequencing data presented here were generated from whole transcriptome sequencing of n=3 organoids per condition.

CRPC-NEPCs exhibit reactivation of EMT plasticity and the acquisition of stem-like cell properties. We, therefore, suspected that, among neuroendocrine tumors, distinct integrin ligands would further upregulate genes to drive an EMT transition. While we observed an enrichment of the EMT pathway in all three matrices, REDV and GFOGER ECMs had a similar signature that was distinct from RGD ligands (**Figure 6B**). Among genes that were near equally enriched in all three ECMs, as compared to Matrigel, cytoskeletal gene vimentin *(VIM)* was significantly upregulated. At the onset of migratory behavior, cells often initiate the expression of vimentin, an intermediate filament protein that forms networks extending from a juxtanuclear cage to the cell periphery. Intrigued by these results, using confocal microscopy, we investigated whether Vimentin was upregulated at the protein level and specifically localized in synthetic organoids as compared to Matrigel. We suspected that CRPC-NEPCs are adapting invasive behavior in synthetic ECMs, and indeed, Vimentin was more expressed in REDV organoids than in Matrigel and localized near the nucleus (**Figure 6C**). Intriguingly, among CRPC-NEPC organoids we uncovered upregulation of CD44, which is an adhesion protein typically associated with a stem-like phenotype, and PLAU1 (**Figure 6D**), which has been shown to promote metastasis and an invasive phenotype in prostate cancer.

We further found that, among CRPC-NEPCs, the ECM receptor pathway was upregulated (**Figure 6B, Supplementary Fig. 10**), indicating that perhaps the cells were responding to the microenvironment through ECM-integrin interactions. Indeed, RGD matrices enriched for *ITGB3* gene, which corroborates with RGD’s ability to bind to integrin αvβ3. To understand what genes were enriched in the epigenetic pathways, we performed a KEGG_Epigenetic analysis and EZH2 target gene analysis (**Figure 6B**). We observed a large number of enriched genes in organoids grown in synthetic ECMs, as compared to Matrigel and among these, a few were differentially regulated within GFOGER, REDV, and RGD. Specifically, we observed differences in the enrichment of *HIC1*, which plays a role in chromatin condensation, with lower enrichment in REDV and GFOGER. Among the EZH2 target genes, *GNA14* was differentially enriched, with negative scores in REDV and GFOGER. GNA14 depletion inhibits the proliferation of cells and potentially slows down growth genes or cell cycle genes. Previous studies have further shown that EZH2 facilitates cell proliferation by repressing cyclin-dependent kinase inhibitors, especially CDKN2A (p16Ink4a p14Arf), which is a canonical Polycomb target gene and tumor suppressor and inhibits progressions from G1 into S phase. CDKN2A was modestly enriched in all three ECM organoids. Overall, we report an increase in genes associated with metastasis and EMT transition, highlighting that these hydrogels may be a useful model for disease development and evolution.

### Synthetic hydrogel organoids identify DRD2 as a novel therapeutic for CRPC-NEPCs

We questioned whether synthetic ECM-grown organoids would identify new therapeutic targets in CRPC-NEPC as there are limited treatment options for CRPC-NEPCs, and as of yet there are no approved targeted therapies. To identify a potential targeted therapy, we hypothesized that differentiation to a neuroendocrine phenotype would result in a greater expression of genes associated with a neural lineage. To validate this hypothesis, we first investigated the patient transcriptomic cohort, seeking to identify genes upregulated in CRPC-NEPC relative to CRPC-Adeno or benign tissues. To identify potential therapeutic targets, we first sought to identify druggable receptors that are upregulated in neuroendocrine tumors. An integrated mRNA analysis in patient tumors revealed that Dopamine Receptor D2 (DRD2) was upregulated in CRPC-NEPC patients compared to CRPC-Adeno patients. Dopamine is a major modulatory neurotransmitter in the central nervous system (CNS) and thus affects neuroendocrine secretion. DRD2 is highly upregulated in neuroendocrine cancers, and DRD2 targeting has emerged as a promising therapy in the treatment of glioma^34^. Neuroendocrine patients among the transcriptomic cohort expressed a significant 27-fold higher expression of DRD2 relative to CRPC-Adeno patients (**Figure 7A**).

**Figure 7.**
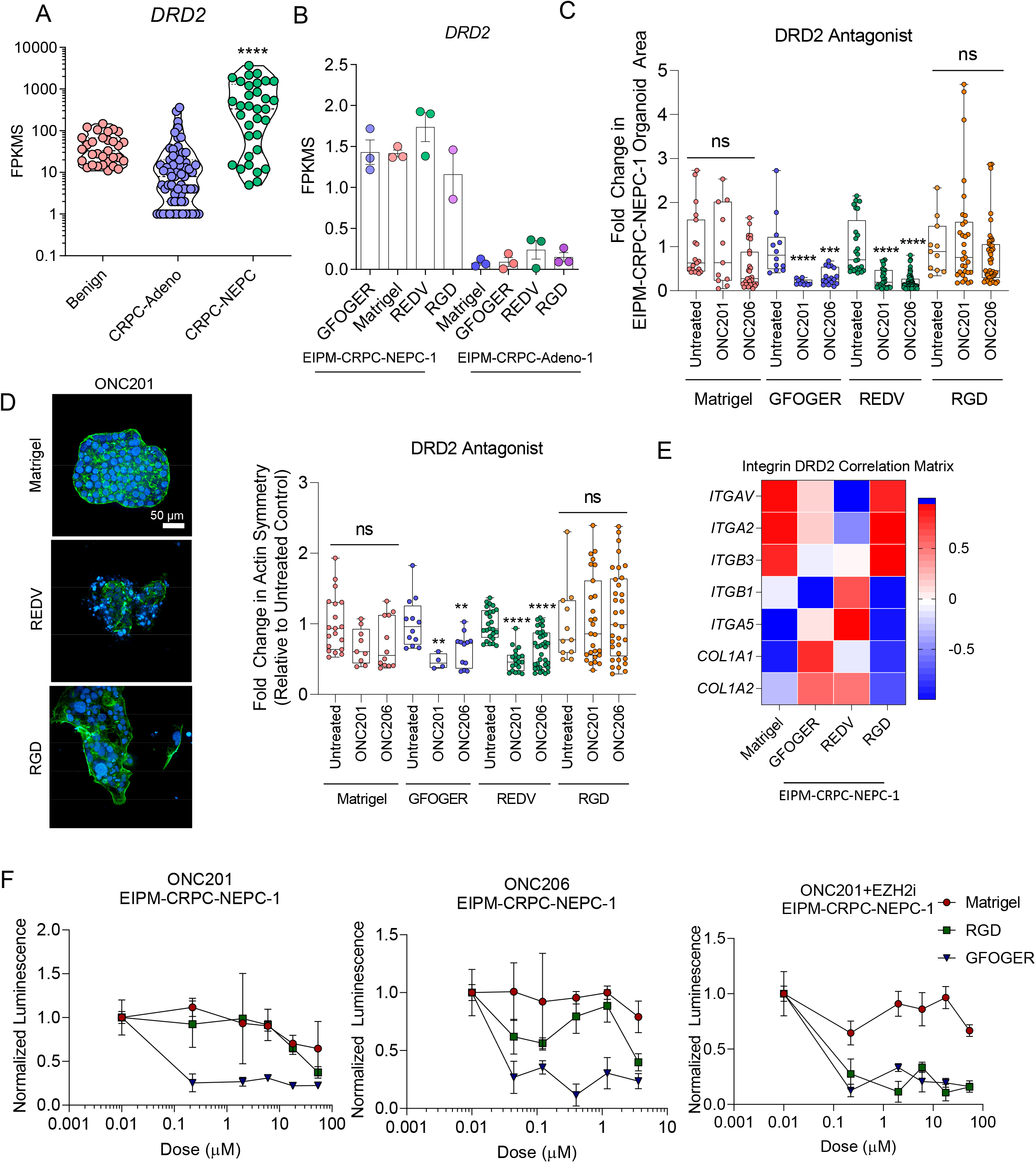
Hydrogel-based organoids identify Dopamine Receptor 2 as a novel therapeutic target in neuroendocrine prostate cancer. **A)** Transcriptomic expression of Dopamine Receptor 2 (DRD2) across disease progression from the WCMC patient cohort (n=31 Benign, n=74 CRPC-Adeno, n=35 CRPC-NEPC). Groups were compared by a one-way ANOVA with a posthoc Tukey’s test with ****p<0.0001. **B)** DRD2 expression in precision and Matrigel prostate organoids (n=3 per condition). **C)** High content imaging of DRD2 inhibition on Matrigel and synthetic-based organoids as evaluated by organoid growth area. Treated groups were compared to untreated controls by a one-way ANOVA, with posthoc Tukey’s test with **p<0.01, ***p<0.001, and ****p<0.0001. **D)** Left, representative confocal images from a high content analysis of ONC201 efficacy, with DAPI in blue and actin in green. Right, quantification of actin symmetry of organoids under DRD2 antagonist treatment. Treated groups were compared to untreated controls by a one-way ANOVA, with posthoc Tukey’s test with **p<0.01, ***p<0.001, and ***p<0.0001. **E)** Correlation heatmap of transcriptomic expression of ECM associated signals (n=3 per condition). **F)** Correlation heatmap of transcriptomic expression of DRD2 signal with genes differentially expressed by synthetic organoids (n=3 per condition). **G)** Drug response curves of DRD2 expression, among Matrigel and precision organoids for ONC201, ONC206, and 0NC201 with an EZH2 inhibitor (n=5 per condition).

We further confirmed that DRD2 was highly expressed among the CRPC-NEPC organoids (**Figure 7B**). In contrast, EIPM-CRPC-Adeno-1 organoids did not express the receptor among any ECM conditions or Matrigel.

As a potential treatment strategy for targeted therapy against DRD2, we tested novel therapeutics, imipridone, which are anti-cancer compounds that possess a three-ring heterocyclic core structure with two substitutable basic amines. We specifically tested two emerging small molecule inhibitors ONC201 and ONC206, developed by Oncoceutics, Inc. ONC201 (benzyl-benzylmethyl-impridone or 1,2,6,7,8,9-hexahydroimidazo[1,2-a]pyrido[3,4-e]pyrimidin-5(4H)-one) is a small molecule discovered through a phenotypic screen for p53-independent inducers of TRAIL-mediated apoptosis—currently in two phase II and one phase I clinical trials for high-grade glioma, with an emphasis in diffuse midline glioma. Using a Bayesian machine-learning approach, we had recently identified that its binding target is DRD2^35^, and the drug has recently been shown in a successful phase I study with solid glioma tumors, which similarly express high DRD2^34^. Unlike other oncology drugs, ONC201 and other imipridones selectively target G protein-coupled receptors (GPCRs) that are dysregulated in cancers. It has additionally shown that it is effective against certain subtypes of glioma for which it is currently in two Phase II clinical trials ^34^, and is currently being tested in metastatic neuroendocrine tumors [ClinicalTrials.gov Identifier: NCT03034200]. Similarly, ONC206 (benzyl-flurobenzyl impridone) is an imipridone with highly potent activity in preclinical models of neuroendocrine tumors. Imipridones have shown high bioavailability as well as a good clinical safety profile and are therefore an encouraging approach for neuroendocrine tumors.

We first performed high content imaging to characterize the morphological response of these organoids to the DRD2 antagonist (**Figure 7C)**. Among EIPM-CRPC-NEPC-1 tumors grown in Matrigel and RGD organoids, we observed no response to drug treatment. However, both REDV and GFOGER-based organoids had a significantly smaller cluster area after treatment with either ONC201 or ONC206. Intriguingly, since DRD2 acts on cytoskeleton inhibition through microtubules^35^, we examined the changes in actin symmetry after the addition of DRD2 antagonist. A comparison of maximum intensity projections yielded that among Matrigel and RGD organoids, cell clusters maintained their actin and cytoskeletal structure **(Figure 7D, left)**. On the other hand, REDV organoids treated with DRD2 antagonists were notably less symmetrical, with a loss of underlying actin structure **(Figure 7D, right)**. We observed a strong correlation between Fibronectin *(FN1)* and DRD2 expression among fibronectin-mimicking REDV organoids **(Figure 7E)**, as determined by RNA sequencing analysis. Similarly, collagen and DRD2 were strongly correlated among GFOGER organoids, suggesting that ECM mimicking peptides may impact the interactions between DRD2 and the corresponding ECM genes. Notably, Matrigel correlations between DRD2 expression and ECM components were weaker, indicating that modular organoids may be required to elucidate these interactions. The ECM correlations were further confirmed with an increased correlation among α5 and β1 integrin subunit in REDV organoids, but not αv and β3 integrins, which correlated more with the RGD organoids **(Figure 7E)**.

We tested whether the ECM matrices could impact the ONC201 and ONC206 in a dose-response manner. Using a metabolic-based luminescent assay, we determined that drug treatment up to supraphysiological doses failed to kill organoids in Matrigel or RGD niches, while GFOGER organoids showed susceptibility to treatment at sub-micromolar doses, among both ONC201 and ONC206 treatment (**Figure 7F**). We next hypothesized that epigenetic reprogramming by EZH2 pre-treatment would modulate the response in an ECM specific manner. When treated with an EZH2 inhibitor for 5 days, RGD organoids became susceptible to DRD2 inhibition, while Matrigel organoids still failed to predict response to ONC201. Taken together, these results highlight the interplay between ECM and synergistic epigenetic targeting in the treatment of CRPC-NEPCs.

It is not well understood whether the components of tumor niche can contribute to the resistance and poor survival observed with CRPC-NEPC relative to other prostate cancer subtypes. In this work, we have engineered synthetic ECM to grow organoids based on patient proteomics and transcriptomics data and characterized these interactions at the phenotypic and transcriptomic level. Phenotypically, we observed that prostate organoids exhibit a spread morphology, reminiscent of epithelial to mesenchymal transition. Concurrently, we observed that transcriptomic data validates a transition into a more mesenchymal phenotype. Using these organoids, we have identified a new potential therapeutic target for CRPC-NEPC and elucidated how ECM-integrin interactions can drive transcriptomic changes, which subsequently renders these tumors susceptible to EZH2 and DRD2 antagonist activity. Synthetic hydrogels can be engineered to recapitulate integrin signaling present *in vivo*, and these hydrogels hold promise across a wide range of tumors. Existing datasets, such as whole transcriptome sequencing, can be leveraged to generate synthetic organoids that can be used to identify novel drug targets. Finally, through identifying ECM-therapeutic target interactions, we identify targeted compounds that would have failed according to the current organoid paradigm. This work will motivate future efforts in designer platforms for prostate cancer, with applications across the fields of cancer biology and precision medicine.

## Data Availability

RNA sequencing datasets will be made available before peer-reviewed publication.

## Data Availability

RNA sequencing datasets will be made available via NCBI and accession codes will be available before publication.

## Author Contributions

Experiments and analyses were performed by MJM, with support from RB, JMB, MA, CC, LP, and MLM. DRD2 inhibitors were provided by VP. The concept was conceived by AS, along with scientific feedback from OE, HB, JMM, and MR. Funding was arranged by H.B, J.M.M, M.R, M.A, O.E., and A.S. The manuscript was written by AS and MJM, and all authors read and provided feedback.

## Acknowledgment

The authors acknowledge financial support from the National Cancer Institute of the US National Institutes of Health (5P50CA211024 awarded to H.B, J.M.M, M.R, M.A, O.E., and A.S.), a US National Science Foundation CAREER award (DMR-1554275 awarded to A.S.), and the Innovative Molecular Analysis Technology program of the US National Cancer Institute (NIH R33-CA212968-01 awarded to A.S.). The authors acknowledge financial support from the NIH Immunoengineering T32 training grant to M.J.M (NIBIB, 1T32EB023860-01A1). Opinions, interpretations, conclusions, and recommendations are those of the authors and are not necessarily endorsed by the funding agency.

## Methods and Materials

### Proteomics analysis

Prostate cancer tissue processing and mass spectrometric analysis were performed according to a published protocol with minor modifications^36^. Briefly, tissues were homogenized, and protein was extracted using 9M urea. After extraction, protein samples were subjected to reduction with dithiothreitol (Sigma) and alkylation with iodoacetamide (Sigma) before overnight digestion with Trypsin (Gibco) at 37 °C. The resulting peptides were desalted and labeled with the tandem mass tag (TMT) reagents according to the manufacturer’s protocol (catalog no. 90110, Thermo Fisher Scientific). Labeled peptides were mixed and desalted before peptide fractionation by high pH reverse phase chromatography to obtain 12 fractions. About 5% of each fraction was desalted and used for protein expression profiling and 95% was used for phosphopeptide enrichment by titanium dioxide (TiO2). These fractions were analyzed by Liquid Chromatography with tandem mass spectrometry (LC-MS). For the LC-MS data acquisition, a Thermo Fisher Scientific EASY-nLC 1200 coupled on-line to a Fusion Lumos mass spectrometer (Thermo Fisher Scientific) was used. Buffer A (0.1% formic acid in water) and buffer B (0.1% formic acid in 80 % acetonitrile) were used as mobile phases for gradient separation. A 75 μm I.D. column (ReproSil-Pur C18-AQ, 3 μm, Dr. Maisch GmbH, German) was packed in-house for peptides separation. Peptides were separated with a gradient of 5–10% buffer B over 1 min, 10%-35% buffer B over 110 min, and 35%-100% B over 10 min at a flow rate of 300 nL/min. The Fusion Lumos mass spectrometer was operated in data-dependent mode. Full MS scans were acquired in the Orbitrap mass analyzer over a range of 400-1500 m/z with resolution 120,000 at m/z 200. Top 15 most abundant precursors were selected with an isolation window of 0.7 Thomson and fragmented by higher-energy collisional dissociation with a normalized collision energy of 40. MS/MS scans were acquired in the Orbitrap mass analyzer. The automatic gain control target value was 1×10^6^ for full scans and 5×10^4^ for MS/MS scans respectively, and the maximum ion injection time is 54 ms for both.

For protein identification, the raw data files were processed using the MaxQuant^37^ computational proteomics platform version 1.6.1.0 (Max Planck Institute, Munich, Germany). The fragmentation spectra were used to search for the UniProt human protein database (downloaded September 21, 2017). Oxidation of methionine and protein N-terminal acetylation were used as variable modifications for database searching. For the phosphopeptide analysis, phosphorylation on serine, threonine, and tyrosine was also used as variable modification. The precursor and fragment mass tolerances were set to 7 and 20 ppm, respectively. Both peptide and protein identifications were filtered at a 1% false discovery rate based on a decoy search using a database with the protein sequences reversed.

### Patient cohort description, pathology classification, and organoid development

Fresh tumor biopsy specimens were obtained and processed into organoids, as outlined in our previous work^13^. Briefly, biopsy specimens were obtained through a next-generation sequencing-based clinical study^38, 39^ approved by the Institutional Review Board at Weill Cornell Medicine (IRB #1305013903). All hematoxylin and eosin-stained and immunohistochemistry slides were reviewed by pathologists (MA and JMM). Histologic criteria were from the proposed classification of prostate cancer with neuroendocrine differentiation^8^.

For tissue processing and development, we again followed the pipeline outlined in our previously established work^13^. We isolated fresh biopsy samples and placed them in DMEM (Invitrogen) supplemented with GlutaMAX (1x, Invitrogen), 100 U/Lml, 100 μg/ml streptomycin (Gibco), Primocin 100 μg/ml (InvivoGen), and 10 μmol/l ROCK inhibitor (Selleck Chemical Inc.). Tissue samples were washed 2x, and the tissue was then enzymatically digested in 250 U/ml collagenase IV (Life Technologies) and TrypLE express (Gibco) in a 1:2 ratio. Incubation time was dependent on the amount of tissue, ranging from 30-90 minutes.

After digestion, tissue fragments were washed in Advanced DMEM/F12 and centrifuged at 300 rcf for 3 min. The pellet was resuspended with prostate-specific culture media composed of Advanced DMEM (Invitrogen) with GlutaMAX (1×, Invitrogen), 100 U/ml penicillin, 100 μg/ml streptomycin (Gibco), Primocin 100 μg/mL (InvitroGen), B27 (Gibco), *N*-Acetylcysteine 1.25 mM (Sigma-Aldrich), Mouse Recombinant EGF 50 ng/ml (Invitrogen), Human Recombinant FGF-10 20 ng/ml (Peprotech), Recombinant Human FGF-basic 1 ng/ml (Peprotech), A-83-01 500 nM (Tocris), SB202190 10 μM (Sigma-Aldrich), Nicotinamide 10 mM (Sigma-Aldrich), (DiHydro) Testosterone 1 nM (Sigma-Aldrich), PGE2 1 μM (R&D Systems), Noggin conditioned media (5%) and R-spondin conditioned media (5%). The final resuspended pellet was combined with Matrigel (Corning) in a 1:2 volume Matrigel, with 6 50 μl droplets pipetted onto each well of a six-well suspension culture plate (Sarstedt LTD). The plate was then incubated for 30 min at 37°C to crosslink Matrigel before 3 ml of media was added to each well. The culture was maintained with fresh media changed twice a week and passages taking place as outlined previously. Throughout prostate organoid development, cultures were screened for various *Mycoplasma* strains using the MycoAlert Kit (Lonza) and confirmed negative before being used for experimental assays.

### RNA Sequencing and analysis of synthetic hydrogel-based organoids

RNA-sequencing and data processing of synthetic organoids was performed according to the protocol described earlier by us^7, 40^. Briefly, RNA was extracted from frozen material for RNA-sequencing (RNA-seq) using the Promega Maxwell 16 MDx instrument, (Maxwell 16 LEV simplyRNA Tissue Kit (cat. # AS1280)). Specimens were prepared for RNA sequencing using TruSeq RNA Library Preparation Kit v2. RNA integrity was verified using the Agilent Bioanalyzer 2100 (Agilent Technologies). cDNA was synthesized from total RNA using Superscript III (Invitrogen). Sequencing was then performed on GAII, HiSeq 2000, or HiSeq 2500^7, 40^. All reads were independently aligned with STAR_2.4.0f1^41^ for sequence alignment against the human genome sequence build hg19, downloaded via the UCSC genome browser [http://hgdownload.soe.ucsc.edu/goldenPath/hg19/bigZips/], and SAMTOOLS v0.1.19^42^ for sorting and indexing reads. Cufflinks (2.0.2)^43^ was used to estimate the expression values (FPKMS), and GENCODE v19^44^ GTF file for annotation. Rstudio (1.0.136) with R (v3.3.2) and ggplot2 (2.2.1) was used for the statistical analysis. The gene counts from htseq-count^45^ and DESeq2 Bioconductor package^46^ were used to identify differentially expressed genes. The hypergeometric test and Gene Set Enrichment Analysis (GSEA)^47^ was used to identify enriched signatures using the *different* pathways collection in the MSigDB database^48, 49^. We used the GSEA pre-ranked method from GSEA for our purpose.

### Nanostring analysis

Molecular characterization was completed on both patient samples and organoids. We utilized a targeted gene panel described previously^50^. Briefly, we utilized a targeted gene panel for this study using the NanoString nCounter that was applied to FFPE, biopsy, and RP tissues with limited RNA input requirement (<300 ng). The gene panel included 163 genes based on known or potential role in prostate cancer progression. Raw data counts were normalized using the nSolver analysis software version 2.0, which normalizes samples according to positive and negative control probe. The edgeR package was used to determine genes that were differentially expressed when comparing the treated and untreated cases. The differentially expressed genes were identified by fitting a generalized linear cluster model, comparing specimens between disease subtypes. Hierarchical clustering in edgeR library package distance was used.

### PEG-4MAL hydrogel organoid

Synthetic organoids were fabricated using 4-armed PEG-4MAL (MW 22,000, Laysan Bio, Inc., >90% purity) and thiolated crosslinkers (DTT:, VPM:. Crosslinkers were obtained from Apptec, including RGD (GRGDSPC), GFOGER (GYGGGP(GPP)_5_GFOGER(GPP)_5_GPC), and REDV (GREDVSPC). PEG-4MAL was dissolved in HEPES buffer at a 20 mM concentration. PEG-4MAL was functionalized with bioadhesive peptides RGD, REDV, or GFOGER (in HEPES at a 10.0 mM concentration to achieve a 5X final ligand density) with at a 4:1 macromer-to-peptide ratio. MMP-degradable (VPM: GCRDVPMSMRGGDRCG) and non-degradable thiolated crosslinkers (DTT) were combined at a defined ratio and a 4:1.5 macromer-to-crosslinker molar ratio. The PEG-4MAL in solution was combined with adhesive peptides and incubated for 30 minutes at 37°C for functionalization.

DU145, EIPM-CRPC-NEPC-1, and EIPM-CRPC-Adeno-1 were isolated from Matrigel, as described earlier by us^13^. After washing 2x in PBS++ (Gibco) solution, prostate cells were suspended in the crosslinker solution. We added PEG-4MAL macromer to the surface of a non-treated plate and added a cell-crosslinker solution at a 1:1 ratio. PEG-4MAL and crosslinker were mixed 5 times to ensure uniform crosslinker. Organoids were crosslinked for 10 min and 37°C. Fresh prostate media was added to the plate and synthetic and Matrigel organoids were cultured before downstream assays.

### Flow cytometry, immunohistochemistry, and microscopy

Tissue sections of patient samples were obtained from the Translational Research Program at Weill Cornell Medicine, previously collected under IRB#1305013903. Immunohistochemistry was performed on deparaffinized formalin-fixed paraffin-embedded sections (organoid, xenograft or patient tissue) using a Bond III automated immunostainer (Leica Microsystems, IL, USA). Heat-mediated antigen retrieval was performed using the Bond Epitope Retrieval solution 1 (ER1) at pH6 or 2 (ER2) at pH9. EZH2 antibody was used at a 1:20 dilution (clone 11/EZH2, BD Biosciences, CA, USA; ER1, 1:20 dilution.)

For flow cytometry analysis, organoids were digested in TrypLE (Gibco) for Matrigel or collagenase (Worthington Biomedical) for PEG-4MAL. After digestion, cells were spun down at 400x to attain single-cell suspensions. Cells were subsequently blocked for half an hour in PBS with 1% BSA (Sigma). Cells were subsequently washed (400x) and resuspended in FACS buffer. After an hour of incubation on ice, cells were washed 3x before being analyzed on an Accuri C6 flow cytometer (Becton Dickinson). Flow cytometry analysis was performed using FlowJo (Treestar). All antibodies or their fluorophore variations were used in these studies at a 1:250 dilution. The complete list of antibodies, clones, and vendors is available in **Supplementary Fig. 14**.

For confocal imaging, cells were seeded onto a glass-bottom coverslip (Matsunami). After culture for seven days to allow organoids to grow, dishes were carefully washed 3X with PBS. Cells were fixed with 4% PFA for 30 min, and organoids were then stained with Phalloidin (40x dilution) for 40 min on ice. Organoids were then permeabilized by incubation with 0.5% Triton x-100 in PBS for 30 min, with a subsequent 3x wash in PBS. Organoids were then incubated with primary antibody for 60 min, washed 3x, and then incubated with secondary antibody for a subsequent 60 min. Finally, organoids were stained with DAPI for ten min. Cells were imaged on an LSM 710 microscope (Zeiss).

For high content imaging, cells were stained as indicated above except on a 96-well glass-bottom plate (X) and imaged using Operetta CLS High-Content Analysis System. Organoid clusters were identified and analysis was performed using Harmony High-Content Imaging and Analysis Software v4.9. Downstream analysis of organoid readouts was performed in Excel and Graphpad Prism 8 (Graphpad). All fluorophores and clones are available in the supplemental data **(Supplementary Fig. 14)**.

### Drug treatment studies

For drug treatment studies, organoids were seeded into either Matrigel or a PEG-4MAL gel. Study timelines were defined by the drug mechanism of action and previous work^13^. For EZH2 studies, organoids were seeded and allowed to grow for 2 days. After 2 days, GSK343 (GlaxoSmithKline) was added at a concentration of 3 μM, according to the previously established EC50 in Matrigel organoids. The organoids were grown further for 5 days and proliferation was quantified using a CellTiterGlo-3D assay (Promega). A longer incubation period (5 days) with an EZH2 inhibitor was necessary to facilitate the epigenetic reprogramming of the cells. Briefly, CellTiterGlo-3D was added to the organoid culture at a 1:1 ratio with media and pipetted to ensure complete mixing. After 30 min, luminescence was evaluated using a Biotek Synergy H4 plate reader.

For DRD2 antagonist studies, cells were similarly seeded and allowed to grow for 5 days. After 5 days, DRD2 antagonists were added according to the denoted concentrations according to a six-fold dilution and incubated for 2 days. This timeline has previously shown efficacy in DRD2 antagonist studies. For luminescence studies, an identical protocol to the EZH2 studies was used. For high content imaging, cells were stained with Actin and DAPI as described above and again imaged on Perkin Elmer Operetta CLS High-Content Analysis System (PerkinElmer). For synergistic studies, organoids were grown for 2 days followed by the addition of EZH2i (GSK343) for 5 days and finally, the DRD2 antagonist was added. Luminescence was quantified after 2 days of DRD antagonist exposure or 2 days of DRD2 antagonist exposure and 5 days of EZH2 inhibitor for synergistic studies.

### Statistical analysis

All statistical analysis was performed using Graphpad Prism 8.0 (Graphpad). Experimental conditions and analysis were compared according to figure captions.

